# Evaluating the consequences of childhood adiposity on the human plasma proteome at three timepoints across the lifecourse

**DOI:** 10.1101/2025.06.20.25329991

**Authors:** Phoebe Dickson, Grace M Power, Tom R Gaunt, George Davey Smith, Tom G Richardson

## Abstract

Applying a lifecourse approach using human genetic data, we sought to evaluate the effect of childhood adiposity on the circulating proteome using measurements taken at three key life stages. We found genetically predicted childhood adiposity to have an effect on 8 proteins measured during childhood (mean age: 9.9 years) including TNFSF11 which is involved in T cell-dependent immune response. Using a mediation framework, we demonstrate that TNFSF11 confers risk of childhood-onset asthma (colocalization posterior probability (PPA)=88.6%) but not adult-onset asthma (PPA=0.2%), highlighting its putative role as a causal intermediate between adiposity and asthma risk specific to early stages in the lifecourse. Additionally, we found that genetically predicted childhood adiposity has an independent effect on 20 proteins measured during midlife after accounting for adulthood adiposity. Replication analyses supported effects of childhood adiposity on CST6 and NOTCH3 which have been found previously to play a role in breast cancer susceptibility.

## Introduction

The human proteome encompasses a rich catalogue of circulating biomarkers which can be explored to help develop improved disease prevention and treatment strategies. Biobank scale plasma proteomic studies now consist of tens of thousands of individuals providing unprecedented statistical power to identify potential proteomic mediators that link modifiable risk factors and disease (1–3). Adiposity in particular represents a major driver of variation across the human proteome, emphasizing the impact that excess weight has on the regulation of circulating proteins (4). However, to date there has not been a comprehensive evaluation of the influence that childhood adiposity may have on the circulating proteome and whether these changes persist throughout the lifecourse into adulthood. This is largely due to the challenges that exist when investigating lifestage-specific effects using conventional epidemiological approaches, given that they can be prone to reverse causation and confounding factors throughout the lifecourse (5).

A previously proposed approach to separate the independent effects of childhood and adulthood adiposity from each other is through the use of human genetics (6). The advantage of this approach is that germline genetic variants are quasi-randomly dispersed throughout a population and typically fixed at birth, making them more robust to confounding and reverse causation compared to conventional approaches (7, 8). We previously identified sets of genetic variants which have differential effects on adiposity during childhood and adulthood timepoints (6). Leveraging these variant sets using an approach known as lifecourse Mendelian randomization (MR) allows their independent effects on disease outcomes to be investigated. For example, we previously found that childhood adiposity increases risk of coronary heart disease (CHD) and type 2 diabetes (T2D), although upon accounting for adulthood adiposity we found that these effects attenuated to the null with limited remnant evidence of an effect (6). This suggests that these childhood effects on CHD and T2D, which are both typically diagnosed during adulthood, are largely explaining by a sustained and long-term influence of being overweight throughout the lifecourse. Additionally, we observed a protective effect of childhood adiposity on breast cancer risk in later life which was found to be independent of adulthood adiposity in our lifecourse model (6, 9).

In this study, we applied lifecourse MR to systematically evaluate whether childhood adiposity has direct and long-term consequences on the circulating human proteome using data measured at three key stages over the lifecourse. These were childhood (mean age 9.9 years, n=2,473) and early adulthood (mean age 24.5 years, n=2,206) using data from the Avon Longitudinal Study of Parents and Children (ALSPAC), and midlife (mean age 56.5 years) using data from up to 34,557 UK Biobank (UKB) participants (2). Replication analyses on proteins highlighted during midlife were undertaken using data obtained from 35,559 Icelanders from the deCODE genetics study (10). For the proteins highlighted by this approach, we conducted systematic downstream analyses using genetic colocalization on disease endpoints to discern whether they may help unravel the biological mechanisms through which childhood adiposity influences lifestage-specific disease risk.

## Results

### Estimating the effect of genetically predicted childhood adiposity on circulating proteins measured during childhood and early adulthood

We constructed a genetic instrument consisting of 313 genetic variants which have previously been shown to robustly associate with childhood body size (6) (Supplementary Table 1). Further details on the derivation and validation of this genetic instrument can be found in the Methods section. In brief, we have previously conducted extensive validation efforts for this instrument using measures of childhood body mass index (BMI) in three independent cohorts; the Avon Longitudinal Study of Parents and Children (ALSPAC) (6), the Young Finns Study (11) and the Trøndelag Health (HUNT) study (12). Furthermore, a recent study found that this genetic instrument more strongly influences DXA-derived total fat mass during childhood compared to lean mass (13), supporting its validity as a genetic proxy for adiposity specifically, rather than simply capturing effects of having a larger body size.

Evaluating the effect of childhood adiposity using this genetic instrument on 92 circulating proteins measured during childhood (mean age: 9.9 years) using data from the ALSPAC cohort identified 8 proteins robust to multiple testing using a false discovery rate (FDR) threshold of <5% (Figure 1, Supplementary Table 2). Amongst these were circulating OPG, which has been implicated in bone density and osteoclast function (14), as well as circulating TNFSF11, which is involved in the regulation of T cell-dependent immune response (15). Additionally, analysing the proteomic data measured during early adulthood from ALSPAC (mean age 24.5 years) provided evidence that genetically predicted childhood adiposity increases levels of three circulating proteins, including inflammatory cytokine interleukin 6 (IL6) (Figure 1, Supplementary Table 3).

**Figure 1.**
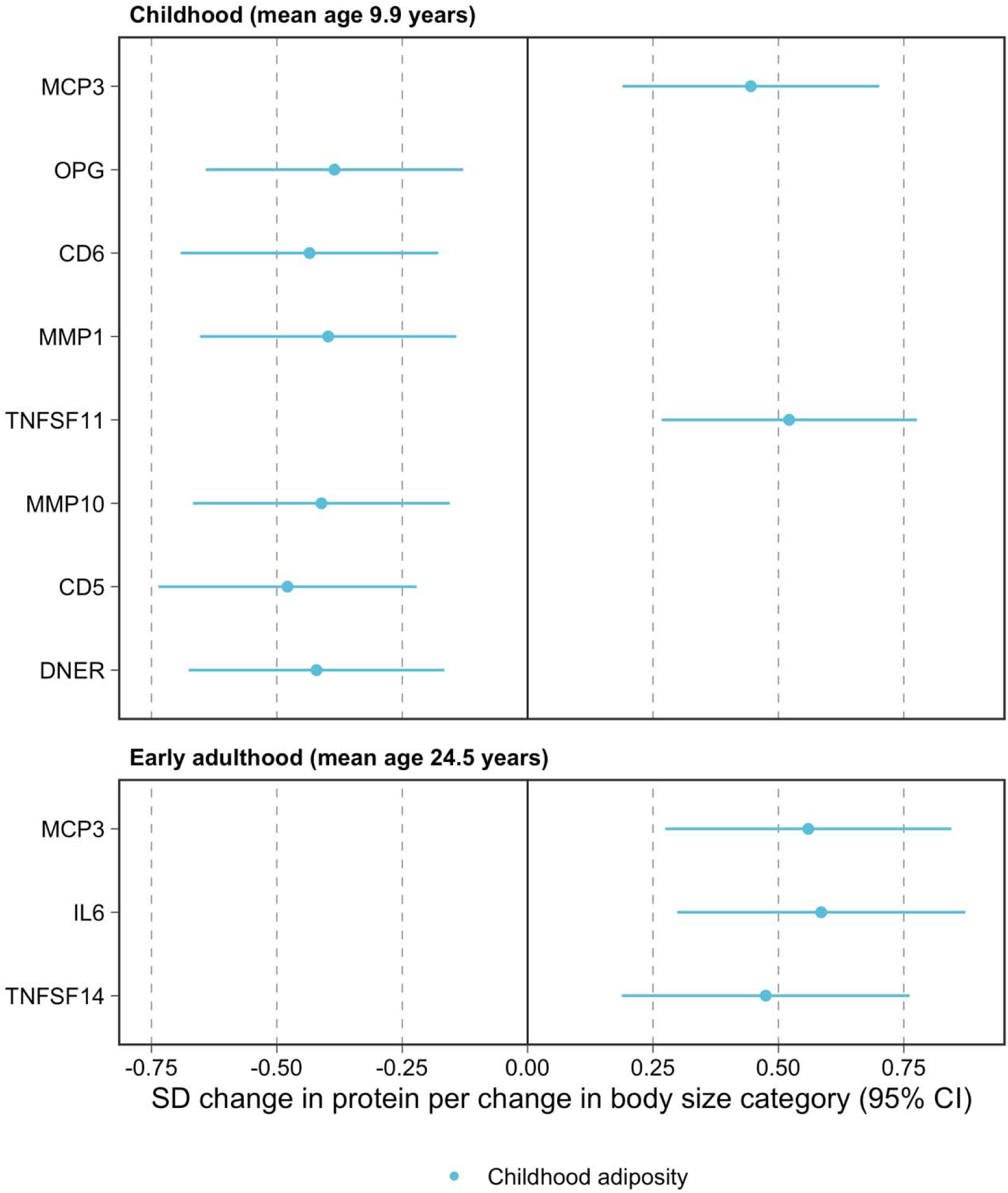
Effects of childhood adiposity on circulating proteins measured during early life in the ALSPAC cohort. Forest plots illustrating the direct effect estimates of childhood (blue) and adulthood (red) adiposity on circulating protein levels measured during A) childhood (mean age 9.9 years) and B) early adulthood (mean age 24.5 years).

### Evaluating the direct and long-term consequences of childhood adiposity on circulating proteins measured during midlife

Applying univariable Mendelian randomization (MR) provided evidence that genetically predicted childhood adiposity influences the levels of 139 circulating proteins measured in the UKB study (mean age 56.5 years) where estimates across 4 different MR methods applied were directionally consistent and robust to FDR<5% corrections (Figure 2A, Supplementary Table 4). Multivariable MR analyses suggested that childhood adiposity has a direct and long-term effect on 20 of these circulating proteins after accounting for genetic effects on adulthood adiposity (Figure 2B, Supplementary Table 5). The remaining proteins highlighted by our univariable analyses may be therefore attributed to the persistence influence of adiposity throughout the lifecourse from childhood into adulthood.

**Figure 2.**
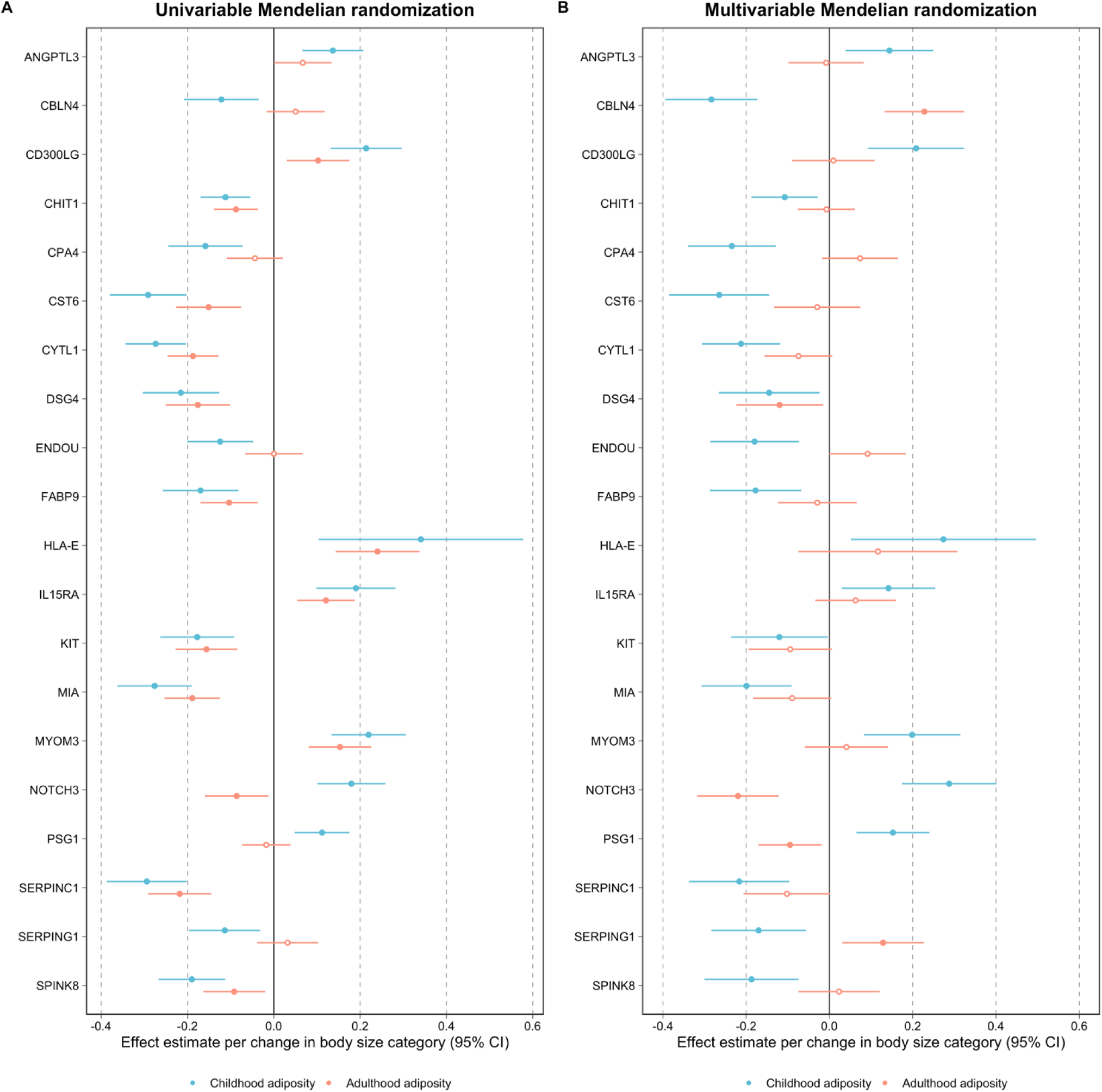
Effects of childhood and adult adiposity on circulating proteins measured during midlife (mean age 56.5 years). Forest plots illustrating the effect estimates of childhood (blue) and adulthood (red) adiposity on circulating protein levels measured during midlife (mean age 56.5) using A) univariable and B) multivariable Mendelian randomization.

In addition, we found evidence of validation for the independent effects of childhood adiposity on circulating NOTCH3, cystatin-M (encoded by CST6) and IL15RA levels amongst the proteins which were also measured using the SomaScan assay in the deCODE proteomic dataset (Supplementary Table 6). Effect estimates were typically very consistent between datasets for these three proteins, for example with a genetically predicted effect of childhood adiposity on elevated levels of circulating NOTCH3 found in the UKB dataset (Beta=0.29 per change in body size category, 95% CI=0.17 to 0.40, P=6.9×10^-7^) and similarly on increased levels of circulating NOTCH3 in the deCODE dataset (Beta=0.29 per change in body size category, 95% CI=0.15 to 0.42, P=2.5×10^-5^).

### Investigating whether circulating proteins influenced by childhood adiposity may act as causal intermediates towards disease risk

Amongst the proteins highlighted by our previous analyses, we found strong evidence that circulating TNFSF11 may play a mediatory role along the causal pathway between childhood adiposity and childhood-onset asthma risk. Figure 3 presents a schematic representation of the approach undertaken to triangulate evidence across three separate analyses alongside their respective supportive results.

**Figure 3.**
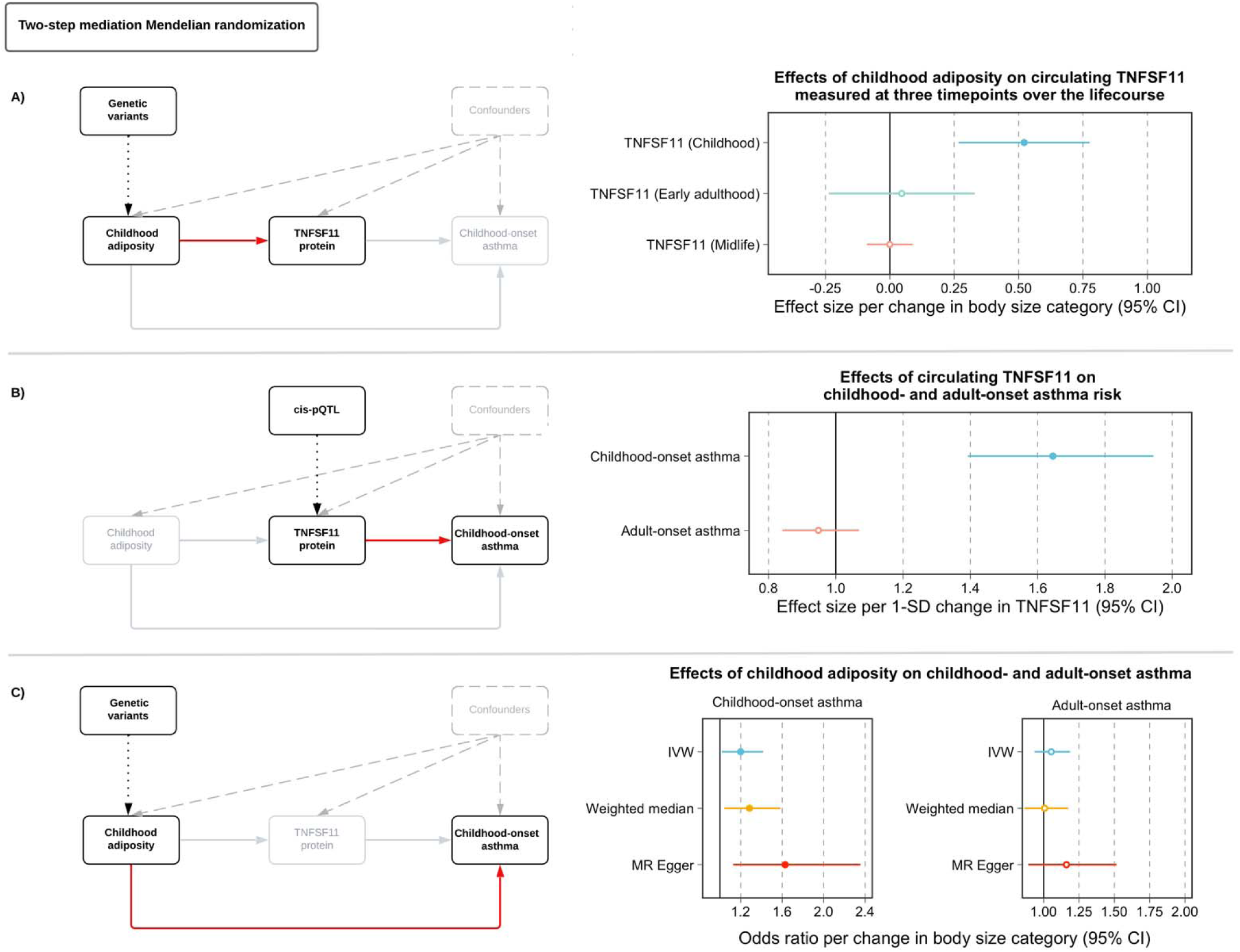
Two-step mediation Mendelian randomization analysis highlights circulating TNFSF11 as mediator between childhood adiposity and childhood-onset asthma risk.

Firstly, we compared effect estimates of genetically predicted childhood adiposity on circulating TNFSF11 across the 3 timepoints where this protein was measured in our study (Figure 3A). We find that childhood adiposity has a strong effect on circulating TNFSF11 levels measured at mean age 9.9 years in the lifecourse (Beta=0.52, 95% CI=0.27 to 0.78, P=6×10^-5^) but markedly weaker evidence based on early adulthood and midlife measures of this protein (early adulthood: Beta=0.05, 95% CI=-0.24 to 0.33, P=0.75, midlife: Beta=-0.0006, 95% CI=-0.09 to 0.09, P=0.99).

Next, using a cis-acting protein quantitative trait loci (pQTL) for TNFSF11 measured at mean age 9.9 years as an instrumental variable we found strong evidence that this protein may play a causal role in childhood-onset asthma risk (OR=1.65 per 1-SD change in protein level, 95% CI=1.39 to 1.94, P=5×10^-9^). In comparison, there was very weak evidence that genetically predicted TNFSF11 measured during childhood has an effect on adult-onset asthma risk (OR=0.95, 95% CI=0.84 to 1.07, P=0.39). Evidence from this analysis was corroborated using genetic colocalization as a complementary analysis, whereby we found strong evidence of a shared causal variant between childhood measured TNFSF11 and childhood-onset asthma risk (posterior probability (PPA)=88.6%) but weak evidence of colocalization between TNFSF11 protein and adult-onset asthma risk (PPA=0.2%) (Figure 4).

**Figure 4.**
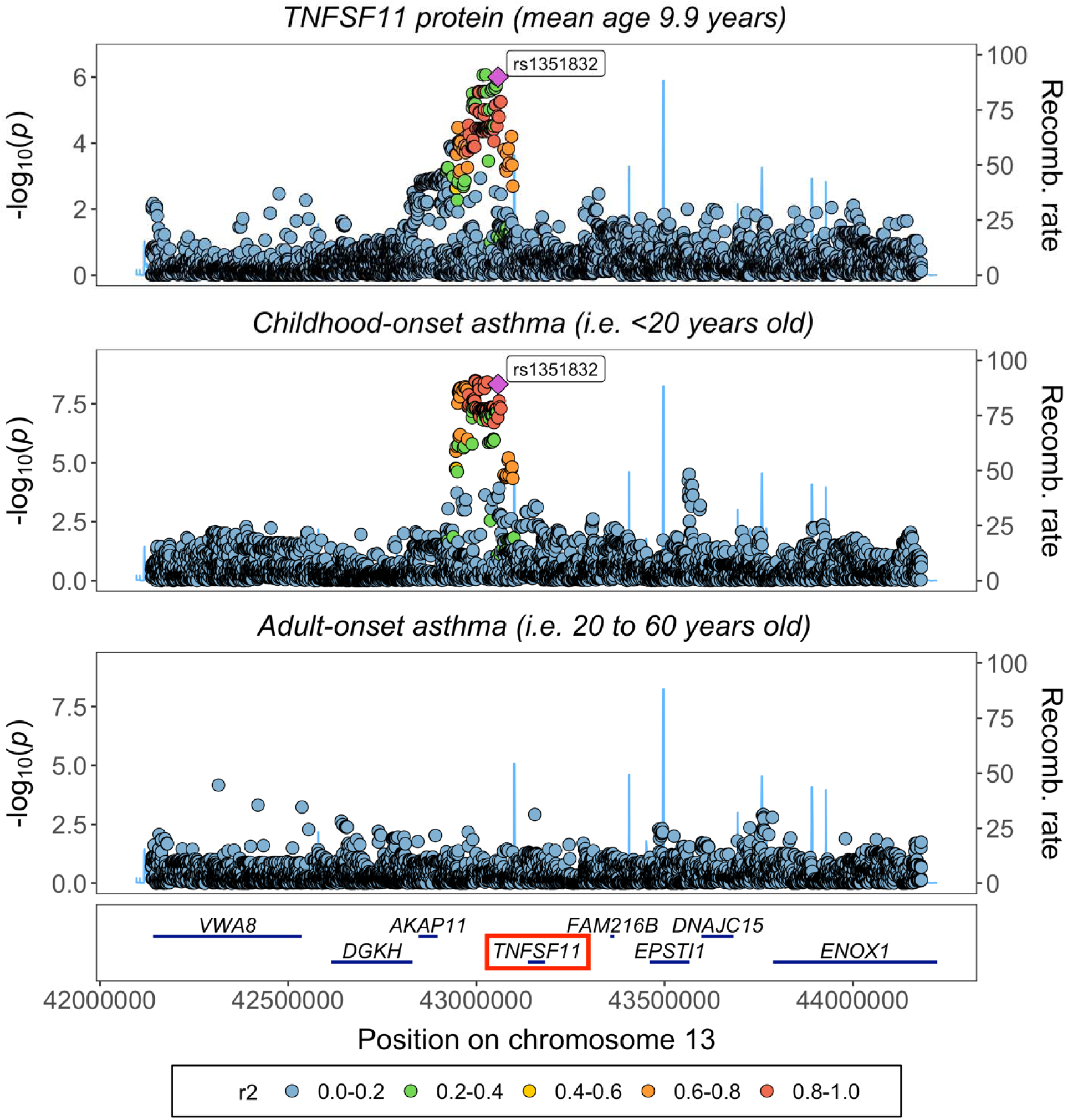
Genetic colocalization between childhood derived circulating TNFSF11 protein with childhood-onset asthma but not adult-onset asthma risk.

Lastly, we conducted a lifecourse MR analysis (as described previously(16)) which provided evidence that genetically predicted childhood adiposity increases risk of childhood-onset asthma (i.e. cases diagnosed at age 19 years or younger) supported by 3 separate MR estimators, whereas there was limited evidence that it has an effect on adult-onset asthma risk (i.e. diagnosed between 20 to 60 years old) (Figure 3C). Full sets of results from our downstream analyses on proteins using genetic colocalization and MR can be found in Supplementary Table 7 and Supplementary Table 8 respectively.

## Discussion

Our findings provide evidence from human genetics that childhood adiposity begins to exert its influence on the circulating proteome as early as age 10 in the lifecourse. In addition, applying a lifecourse approach provided evidence that genetically predicted childhood adiposity has a long-term effect on the levels of 20 proteins measured during midlife (i.e. mean age 56.5 years old) even after accounting for adulthood adiposity in our model. Taken together, the proteins highlighted in this study, either as early life alterations or putative lifelong effects, should be prioritised by future research to better understand their mechanistic role in disease aetiology and whether they may serve as valuable biomarkers for disease prevention.

Amongst the 8 circulating proteins measured at mean age 9.9 years in the lifecourse whose levels were robustly influenced by childhood adiposity, the most compelling evidence that these early life alterations may have downstream consequences for disease risk was found for TNFSF11. This protein, also referred to commonly as RANKL, has been long established to play an important role in the immune system (15), initially discovered as a T-cell derived activator of dendritic cells (17). Across the three timepoints assessed in this study, we found that genetically predicted childhood adiposity only had a strong effect on circulating TNFSF11 during the childhood timepoint (mean age 9.9 years). Furthermore, applying a mediation framework highlighted a causal role for TNFSF11 protein on risk of childhood-onset asthma (i.e. diagnosed at 19 years or earlier in the lifecourse) but very limited evidence that this protein may confer risk of asthma during adulthood. Taken together, our findings suggested there may be a critical window during the early stages of the lifecourse where TNFSF11 acts as a causal intermediate along the pathway between childhood adiposity and risk of childhood-onset asthma.

From the analysis of proteins measured during early adulthood (i.e. mean age 24.5 years), three survived corrections for multiple testing including circulating interleukin-6 (IL6). IL6R blockade is currently being pursued as a potential therapeutic strategy for the treatment of cardiovascular and immune-related endpoints. Previous Mendelian randomization studies have provided supportive evidence of this for outcome such as coronary heart disease (18), ischemic stroke (19) and sepsis (20). Novel insight from this current study suggests that the previously observed effect of adiposity on circulating levels of IL6 may begin to manifest during early adulthood.

Amongst the 20 proteins whose levels were influenced by childhood adiposity independent of adulthood adiposity included circulating NOTCH3, CST6 and IL15RA which are noteworthy given that they provided evidence of replication in the deCODE study measured using a different technology. Recent evidence from breast cancer cell lines has found that NOTCH3 may inhibit tumorigenesis and the emergence of breast cancer through the transactivation of PTEN in breast cancer cell lines (21). Our findings suggest that childhood adiposity increases levels of circulating NOTCH3 measured in adulthood. This result is therefore directionally consistent with the possible role of NOTCH3 as a mediator of the protective effect of childhood adiposity on breast cancer risk, which to date remains unexplained (22). Future work should therefore focus on investigating the proportion of a potential mediatory role that NOTCH3 may explain along this causal pathway, as well as whether this effect is independent of other proposed mediators such as mammographic density (9). Similar investigations should be conducted for circulating cystatin-M encoded by the CST6 gene, which has been found to be down-regulated in metastatic breast tumour cells compared to primary tumour cells (23), as well as cytokine receptor IL15RA which has been previously implicated in immunoregulatory functions (24).

An important consideration when interpreting the findings of this study is that, whilst analysing plasma circulating proteins provides exceptional statistical power given the large sample sizes of whole blood-derived measures, this comes with the caveat that effects confined to more biologically relevant tissues and cell types may be overlooked. Further research into how representative the effects elucidated in this research are across human tissues should yield further insight into the novel biology underlying these findings. Additionally, it should be noted that our genetic instrument for childhood adiposity is based on recall data, which is why we sought its validation in 3 cohorts independent of UK Biobank and additionally found it to be more strongly predictive of measured childhood body mass index and DXA-assessed adiposity measures in comparison to the adulthood adiposity genetic score (6, 11–13).

In conclusion, our findings emphasise the value of using human genetics to separate the effects of childhood and adulthood risk factors on molecular traits such as circulating proteins. In particular, the role of TNFSF11 acting as a putative mediator for the effect of childhood adiposity on childhood-onset asthma risk serves as an important proof of concept that investigating lifecourse-specific measures of circulating proteins, together with age-of-onset information for disease endpoints, can help elucidate time-varying effects which may indicate windows of opportunity for intervention throughout the lifecourse.

## Methods

### Instruments for lifecourse Mendelian randomization

Derivation of genetic instruments for childhood and adult adiposity have been described in detail previously (6). In brief, genome-wide association studies (GWAS) were conducted on 453,169 UKB participants who reported whether they were ‘thinner’, ‘about average’ or ‘plumper’ at age 10 years compared to the average. GWAS on the same sample were conducted based on their measured adult body mass index (mean age: 56.5 years old) which was categorised into the same proportions as the childhood adiposity variable for comparative purposes. In total, there were 313 independent instruments for childhood adiposity and 580 independent instruments for adult adiposity based on genome-wide corrections (i.e. P<5×10^-8^). Amongst these, 92 of the childhood instruments were also associated with adulthood adiposity (29.3%) and 87 of the adult instruments were associated with childhood adiposity (15%) based on genome-wide corrections. A full list of genetic instruments used in this study can be found in Supplementary Tables 1 & 9. All individual participant data used in this study were obtained from the UK Biobank (UKB) study, who have obtained ethics approval from the Research Ethics Committee (REC; approval number: 11/NW/0382) and informed consent from all participants enrolled in UKB.

These instruments have been extensively validated in three independent cohorts; the Avon Longitudinal Study of Parents and Children (ALSPAC) (6), the Young Finns Study (11) and the Trøndelag Health (HUNT) study (12). Furthermore, a recent study found that these childhood genetic instruments more strongly influence DXA-derived total fat mass during childhood compared to lean mass (13), supporting their validity as genetic proxies for adiposity specifically, rather than simply capturing effects of having a larger body size.

### Childhood and early adulthood measures of plasma proteins from the ALSPAC cohort

The Avon Longitudinal Study of Parents and Children (ALSPAC) is a population-based cohort investigating genetic and environmental factors that affect the health and development of children. The study methods are described in detail elsewhere (25, 26). In brief, 14,541 pregnant women residents in the former region of Avon, UK, with an expected delivery date between April 1, 1991 and December 31, 1992, were eligible to take part in ALSPAC. Detailed phenotypic information, biological samples and genetic data which have been collected from the ALSPAC participants are available through a searchable data dictionary and variable search tool (http://www.bris.ac.uk/alspac/researchers/our-data/). Consent for biological samples has been collected in accordance with the Human Tissue Act (2004). Written informed consent was obtained for all study participants. Ethical approval for this study was obtained from the ALSPAC Ethics and Law Committee and the Local Research Ethics Committees.

Detailed methods on Olink derived protein measures in ALSPAC have been described previously (27). In brief, heparin-stored plasma samples derived from ALSPAC participants at mean ages 9.9 years (range: 8.9 to 11.5) and 24.5 (range: 22.4 to 26.5) years were analysed using the Olink Target 96 platform (27). After matching on genotyping data and covariates, final sample sizes for age 9 analyses were up to n=2,473 and age 24 analyses were up to n=2,206.

### Midlife measures of plasma proteins from the UK Biobank and DeCODE cohorts

Data on circulating proteins were obtained from a previously conducted GWAS of n=34,557 UKB participants of European descent conducted by the UKB Pharma Proteomics Project (2). A total of 2,941 proteomic measures were generated from whole blood derived samples randomly selected from the baseline cohort of UKB (mean age: 56.5 years, range: 39 to 73) using the antibody-based Olink Explore 3072 platform. For proteins identified in the UKB datasets as robustly influenced by childhood adiposity based on criteria described in our Statistical analysis section, we sought replication of their effects using a GWAS of human proteins quantified with the SomaScan multiplex aptamer assay (version 4) using samples derived from 35,559 Icelanders from the deCODE genetics study (mean age: 55 years) (10). Full technical details on these proteomic datasets can be found in the corresponding publications by Sun et al (2) and Ferkingstad et al(10).

### Complex traits and disease outcomes

For any circulating proteins robustly influenced by childhood adiposity (see Statistical analysis section for details), we conducted further genetic analyses to discern whether they may play a role in disease risk and complex trait variation. Outcomes were selected based on previously conducted lifecourse MR studies which provided evidence of an effect of childhood adiposity independent of adulthood adiposity. These were childhood-onset asthma (16), breast cancer (BrCa) (6), bone mineral density (28), type 1 diabetes (T1D) (29) (based on cases with a mean age of 16.5 years age of diagnosis) and heart structure traits (30, 31). All relevant study characteristics and data sources for these outcome datasets can be found in Supplementary Table 10 (32–37).

### Statistical analysis

Analyses conducted in ALSPAC were based on individual-level data whereby univariable and multivariable MR models were applied in turn to evaluate the direct and indirect effect of childhood adiposity on each of the Olink Target 96 measured proteins. Genetic risk scores (GRS) for both childhood and adulthood adiposity genetic instruments were constructed as the sum of the number of adiposity-increasing alleles weighted by the estimates of their effect on adiposity. Univariable MR analyses for the childhood adiposity GRS were adjusted for age, sex and the top 10 principal components in ALSPAC. Any genetically predicted effects robust to a false discovery rate (FDR)<5% on proteins measured at either the childhood or early adulthood timepoints were then taken forward to multivariable analyses. Multivariable MR involved simultaneously fitting both childhood and adulthood adiposity GRS in our model meaning that when childhood adiposity was the primary exposure, adulthood adiposity was controlled for, and vice versa. The same covariates were accounted for as in the univariable analysis.

To systematically evaluate the total effect of childhood adiposity on each of the 2,941 circulating proteins measured during adulthood from UKB, we applied a two-sample MR framework firstly using the inverse variance weighted (IVW) method, which takes SNP-outcome estimates and regresses them on the SNP-exposure associations (38). All genetically predicted effects which were robust to an FDR<5% correction were subsequently analysed using several other approaches which are typically less prone to horizontal pleiotropy than the IVW method. Horizontal pleiotropy is the phenomenon whereby genetic variants influence multiple traits or disease outcomes via independent biological pathways. The additional robust methods used were the weighted median (39), weighted mode (40) and MR-Egger estimators (41).

Additionally, we excluded any instruments which were located at the encoding gene region (i.e. hg38 gene coordinates +/- 1Mb) for their corresponding protein in all MR analyses in this study when proteins were analysed as an outcome. This was to remove any instruments which were potentially prone to reverse causation (i.e. proteins influencing childhood adiposity) given that they were located at the gene region encoding the circulating protein being investigated. Given that circulating proteins were measured during adulthood, it is implausible that they may influence childhood adiposity given that this measure is taken at an earlier stage in the lifecourse. Nonetheless, as an additional sensitivity analysis we also conducted reverse MR analyses by taking independent protein quantitative trait loci located at encoding gene regions (i.e. cis-pQTL) as instruments to evaluate the genetically predicted effect of proteins on childhood adiposity.

For all effects of childhood adiposity on proteins which were directionally concordant and robust to FDR<5% corrections across all 4 methods, as well as providing an MR-Egger intercept term which did not indicate evidence of horizontal pleiotropy (i.e. MR-Egger intercept P>0.05), we lastly applied multivariable MR to evaluate whether effects were independent of adulthood adiposity. This involved using the IVW method to model both childhood and adulthood adiposity simultaneously to estimate the direct effect of childhood adiposity on protein measures after accounting for the effect of adulthood adiposity (42). We again applied an FDR<5% correction to evaluate whether childhood adiposity has a long-term influence on circulating proteins independently of adult adiposity.

Finally, we investigated whether the circulating proteins highlighted from these analyses also provided genetic evidence of an effect on the eight downstream traits and disease outcomes mentioned in the ‘complex traits and disease outcomes section’ which have previous evidence of being influence by childhood adiposity. This was undertaken by 1) evaluating the genetically predicted effects of each protein on the eight outcomes using cis-pQTL instruments and 2) assessing whether there was evidence of genetic colocalization at each protein’s encoding gene region between their circulating protein product and each outcome in turn using the coloc-SuSiE method (43). A posterior probability (PPA) greater than 0.80 was used as a threshold to indicate formal evidence of genetic colocalization. Proteins encoded by genes located in the MHC region of the genome were excluded from these analyses due to the known complex LD structure which exists at this region of the genome.

MR analyses were conducted using the ‘TwoSampleMR’ R package (44) and genetic colocalization analyses were undertaken using the ‘coloc’ R package (43). Forest and locuszoom plots in this paper were generated using the R packages ‘ggplot2’ and ‘gassocplot’ respectively.

## Materials and correspondence

This publication is the work of the authors and TGR will serve as guarantor for the contents of this paper.

## Data Availability

All data referred to in this manuscript is either publicly available or can be accessed via an approved applcation to the UK Biobank or ALSPAC cohorts

## Acknowledgements

We are extremely grateful to all the families who took part in this study, the midwives for their help in recruiting them, and the whole ALSPAC team, which includes interviewers, computer and laboratory technicians, clerical workers, research scientists, volunteers, managers, receptionists and nurses. This research was conducted at the NIHR Biomedical Research Centre at the University Hospitals Bristol NHS Foundation Trust and the University of Bristol. The views expressed in this publication are those of the author(s) and not necessarily those of the NHS, the National Institute for Health Research or the Department of Health. The UK Medical Research Council and Wellcome (Grant ref: 217065/Z/19/Z) and the University of Bristol provide core support for ALSPAC. This publication is the work of the authors and TGR will serve as guarantors for the contents of this paper. Genomewide genotyping data was generated by Sample Logistics and Genotyping Facilities at Wellcome Sanger Institute and LabCorp (Laboratory Corporation of America) using support from 23andMe. We are grateful to the researchers who made their summary-level data available for the purposes of this work and the participants of both the UK Biobank and deCODE genetics studies.

